# Improvement in Delivery of Ischemic Stroke Treatments but Stagnation of Clinical Outcomes in Young Adults in South Korea

**DOI:** 10.1101/2023.04.17.23288712

**Authors:** Jonguk Kim, Jun Yup Kim, Jihoon Kang, Beom Joon Kim, Moon-Ku Han, Jeong-Yoon Lee, Tai Hwan Park, Ji Sung Lee, Keon-Joo Lee, Joon-Tae Kim, Kang-Ho Choi, Jong-Moo Park, Kyusik Kang, Soo Joo Lee, Jae Guk Kim, Jae-Kwan Cha, Dae-Hyun Kim, Kyungbok Lee, Jun Lee, Keun-Sik Hong, Yong-Jin Cho, Hong-Kyun Park, Byung-Chul Lee, Kyung-Ho Yu, Mi-Sun Oh, Dong-Eog Kim, Wi-Sun Ryu, Jay Chol Choi, Jee-Hyun Kwon, Wook-Joo Kim, Dong-Ick Shin, Kyu Sun Yum, Sung Il Sohn, Jeong-ho Hong, Sang-Hwa Lee, Juneyoung Lee, Philip B. Gorelick, Hee-Joon Bae

## Abstract

**Background:** There is limited information on the delivery of acute stroke therapies and secondary preventive measures and clinical outcomes over time in young adults with acute ischemic stroke (AIS). This study investigated whether advances in these treatments improved outcomes in this population.

**Methods:** Using a prospective multicenter stroke registry in Korea, young adults (aged 18–50 years) with AIS hospitalized between 2008 and 2019 were identified. The observation period was divided into four epochs: 2008–2010, 2011–2013, 2014–2016, and 2017–2019. Secular trends for patient characteristics, treatments, and outcomes were analyzed.

**Results:** A total of 7,050 eligible patients (mean age 43.1; men 71.9%) were registered. The mean age decreased from 43.6 to 42.9 years (P_trend_=0.01). Current smoking decreased, whereas obesity increased. Other risk factors remained unchanged. Intravenous thrombolysis and mechanical thrombectomy rates increased over time from 2008–2010 to 2017–2019 (9.5% to 13.8% and 3.2% to 9.2%, respectively; P_trend_’s<0.01). Door-to-needle time improved (P_trend_<.001), but onset-to-door and door-to-puncture time remained constant. Secondary prevention including the administration of dual antiplatelets for noncardioembolic minor stroke (26.7% to 47.0%), direct oral anticoagulants for atrial fibrillation (0.0% to 56.2%), and statins for large artery atherosclerosis (76.1% to 95.3%) increased (P_trend_’s<0.01). Outcome data were available from 2011. One-year mortality (2.5% in 2011–2013 and 2.3% in 2017–2019) and 3-month modified Rankin scale scores 0–1 (68.3% to 69.1%) and 0–2 (87.6% to 86.2%) remained unchanged. The one-year stroke recurrence rate increased (4.1% to 5.5%, P_trend_=0.04), altough the differnce was not significant after adjusting for sex and age.

**Conclusion:** Improvements in the delivery of acute stroke treatments did not necessarily lead to better outcomes in young adults with AIS over the past decade, indicating a need for further progress.

## Introduction

Young adults, commonly defined as individuals aged 18 to 50, constitute 10–15% of the acute ischemic stroke (AIS) population.^1^ Despite a reduction in the incidence of ischemic stroke in high-income countries, there has been a worrisome increase among young adults, particularly in developed countries.^2^ Additionally, healthcare costs for young adults with stroke are approximately 1.6 times higher than for older patients.^3^ Therefore, improving outcomes in young adults with stroke is important, especially given the potential socioeconomic impact due to their longer life expectancy.

However, there have been concerns that young adults with ischemic stroke may not receive optimal treatment. Young patients were more likely to be unaware of risk factors prior to experiencing ischemic stroke,^4^ to present at the hospital later after stroke onset,^5^ and to receive thrombolysis later.^6^ Another issue is the lack of specific treatment guidelines for young patients, despite the differing risk factors and causes compared to older patients.^7, 8^

Over the past 15 years, substantial advancements have been made in acute reperfusion therapy and secondary preventive treatment for ischemic stroke.^9–12^ However, the real-world impact of these advancements on clinical outcomes, particularly in young patients, has not been thoroughly explored for more recent period since the early 2010s. While stroke in young adults is gaining increasing attention, data regarding the impact of recent treatment advances on this population remain limited. Prior investigations have often been constrained by cross-sectional designs,^13, 14^ small sample sizes,^15, 16^ or timing that predated the introduction of more advanced treatments.^13, 16, 17^

In this study, we aim to fill these gaps by examining trends in the application of advanced treatments and the subsequent clinical outcomes for young stroke patients from 2011 to 2019 using data from a multicenter stroke registry in South Korea.

## Methods

### Study Population

This retrospective analysis was based on data from a prospective, multicenter stroke registry - the Clinical Research Collaboration for Stroke in Korea-National Institutes of Health (CRCS-K-NIH) registry. The CRCS-K-NIH is a web-based registry implemented in 2008 to register patients with acute stroke or transient ischemic attack admitted to the 17 participating stroke centers in South Korea. Detailed information on the CRCS-K-NIH registry is provided elsewhere.^18, 19^ The following inclusion criteria applied: age 18–50 years at presentation, diagnosis of AIS defined as the presence of ischemic symptoms for over 24 h or a relevant ischemic lesion on diffusion-weighted MRI, arrival within seven days from when the stroke symptoms were recognized by the patient (first abnormal time [FAT]), and hospitalization between April 2008 and December 2019.

### Ethics Statement

The local institutional review boards (IRBs) of all participating centers of the CRCS-K-NIH registry approved the collection of clinical data to ‘monitor and improve the quality and outcomes of stroke care.’ Given the de-identified nature of the data and the minimal risk to the participants, the necessity for informed consent was waved. Furthermore, approval to use the registry database was secured from all necessary IRBs (B-2112-728-102).

### Data Collection

Information on demographics, risk factors, blood pressure, laboratory data, premorbid disability (modified Rankin scale [mRS]), initial National Institute of Health Stroke Scale (NIHSS), stroke subtype according to the Trial of Org 10172 in Acute Stroke Treatment (TOAST) criteria with some modifications,^20^ FAT, last known well time (LKW), diagnostic evaluations, reperfusion therapies, door-to-needle time (DNT), door-to-puncture time (DPT), and discharge medications were obtained from the registry database. Risk factors were dichotomized into two categories: “newly diagnosed” and “previously known or aware,” and the “aware” category was further divided into “on medication” and “not on medication” based on the treatment status.

As part of a program aimed at monitoring and improving the quality of stroke care and outcomes, functional outcomes, and occurrence of clinical events, including recurrent stroke and death, were prospectively captured at discharge, 3 months, and 1 year after the index stroke through a structured telephone interview conducted by experienced nurse coordinators or a direct interview by treating physicians at outpatient clinics according to predetermined protocols.

Functional outcomes were evaluated using the mRS and were divided into two categories: mRS scores of 0–1 vs. 2–6 and 0–2 vs. 3–6, or treated as an ordinal variable. We also analyzed the occurrence of clinical events, including the time from index stroke onset to recurrent stroke and to death, respectively. Regarding these time-to-event data, patients were censored if they were lost to follow-up or at 1.5 years from the onset of stroke. The operational definitions of all variables are listed in Table S1.

### Statistics

We described demographics, risk factors, stroke characteristics, diagnostic evaluations, and treatments across four epochs: 2008–2010, 2011–2013, 2014–2016, and 2017–2019. For trend analyses, the ANOVA linear contrast test was applied to continuous variables, and the Cochrane–Armitage test was applied to categorical variables. The Mantel– Haenszel chi-square test was used to treat the 3-month mRS as an ordinal variable. The Kaplan–Meier product limit method was implemented to estimate mortality and cumulative incidence of recurrent strokes, which were subsequently compared across the epochs using a log-rank linear contrast test. Among the 63 variables used in the study, 20 variables were found to have one or more missing values, resulting in an overall missing rate of 0.49%. These missing values were removed pairwise during the tabulation and analysis.

Stroke outcomes were analyzed with and without adjustment. Variables for adjustment were predetermined as follows: sex and age for all outcome (Model 1), sex, age, and initial NIHSS score for 3-month mRS score and mortality, and sex, age, stroke risk factors, and stroke subtypes for recurrent stroke (Model 2). For trend analyses in multivariable models, a linear contrast test was applied to binary outcomes in logistic regression models, to survival outcomes in Cox proportional hazard models, and to the whole scale of 3-month mRS scores in an ordinal logistic regression model. Secular trends were analyzed according to age (18–30, 31–40, and 41–50 years) and sex subgroups and presented if needed. Furthermore, secular trends in treatments and outcomes were demonstrated in subgroups of potential contributing factors.

A two-tailed *p*-value of <0.05 was considered statistically significant. All analyses were performed using SAS version 9.4 (SAS Institute Inc., Cary, NC, USA) and R software version 4.0.2 (R Foundation for Statistical Computing, Vienna, Austria).

## Results

### Demographics, Risk Factors, and Subtypes of Ischemic Stroke (Table 1)

Of 70,567 patients (age 68.3±13.0 years; men 58.5%) registered with AIS between 2008 and 2019, 7,050 participants aged 18–50 (10.0%) were selected. Over the 12 years, the proportion of men decreased slightly from 74% to 71% (*p*_trend_=0.10), but it remained more than twice that of women. The mean age decreased in both sexes, although this change was statistically significant only in women. The decrease in mean age in women was attributable to an increase in the proportion of women aged 18–30 (Figure 1).

**Figure 1.**
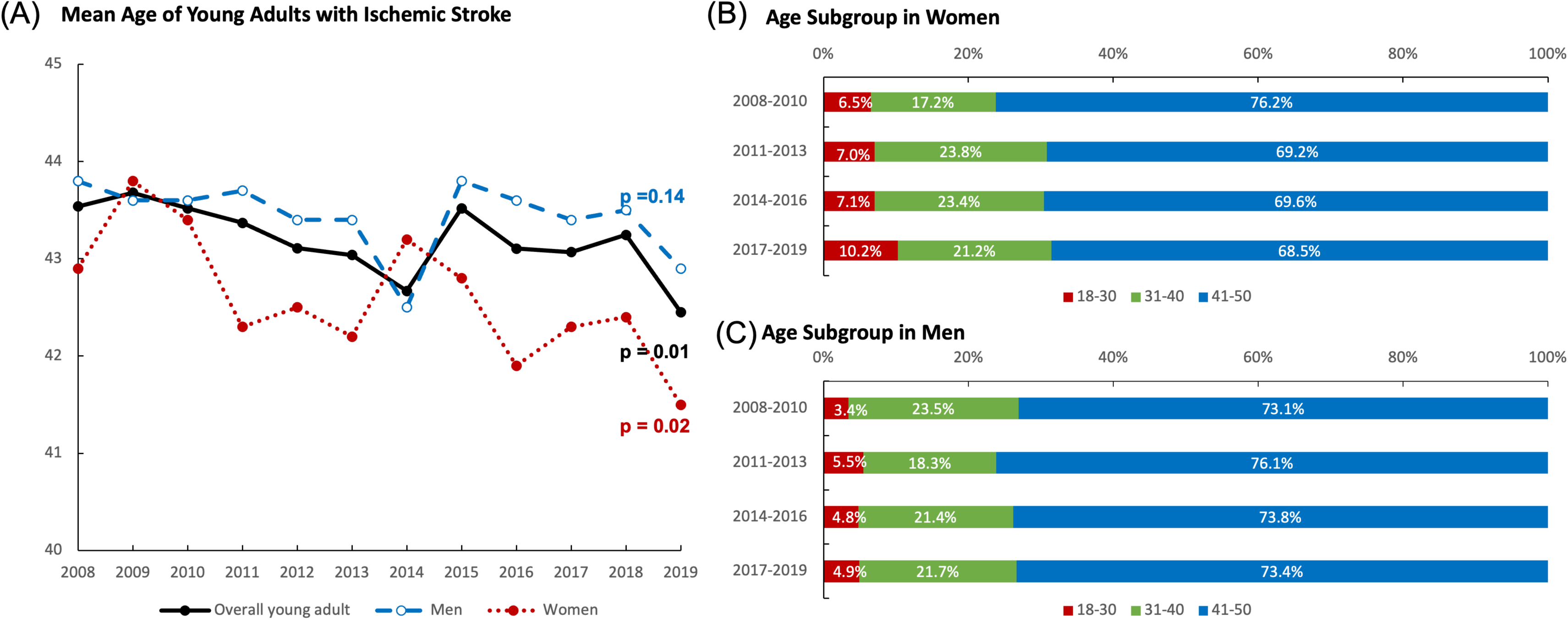
Trends in Mean Age Between 2008 and 2019 (A) Mean age according to sex and calendar year, (B) secular changes in age group proportions in women, and (C) secular changes in age group proportions in men.

Regarding traditional vascular risk factors, we observed significant trends in smoking and obesity. Current smoking decreased by 13.5% in men but increased by 2.5% in women. Obesity (body mass index ≥30 kg/m^2^) more than doubled in men (5.2% in 2008-10 to 13.1% in 2017-19). The increase was somewhat less dramatic in women (5.4% to 9.2%). However, no significant trends were observed regarding hypertension, diabetes mellitus, and hyperlipidemia. Awareness and treatment rates of hypertension, diabetes, and hyperlipidemia before the index stroke remained unchanged except for increased awareness of hyperlipidemia.

Regarding ischemic stroke subtypes, large artery atherothrombosis (LAA) was the most common etiology, followed by small vessel occlusion (SVO) or undetermined etiology (UDE; depending on the epoch). Notably, other determined etiology (ODE) increased by 9.0%, but LAA, SVO, and cardioembolic stroke (CES) decreased by 2.5%, 5.1%, and 4.3%, respectively. Among ODEs, arterial dissection was the most common, followed by intrinsic diseases of the arterial wall, such as Moyamoya disease and fibromuscular dysplasia (Table S2). Arterial dissection significantly increased in both men and women by 4.7% and 5.1%, respectively. The proportion of UDEs remained unchanged throughout the study period.

### Clinical Presentation and Treatments (Tables 2)

Over the 12 years, the onset-to-door time (ODT) remained unchanged; the median FAT-to-door ranged between 8.0 and 9.4h, and the median LKW-to-door between 11.1 and 12.5h. Overall, the proportion of patients hospitalized within 3.5h from LKW was 27.5%. However, initial stroke severity decreased significantly, with a reduction in the mean NIHSS score and the proportion of severe strokes (NIHSS score 16–42) by 0.8 points and 1.9%, respectively.

The IVT rate increased by 3.8% between the first two epochs but plateaued afterward. The DNT followed a similar pattern. The overall increase in IVT rate may be attributed to the rise in IVT rate in patients arriving between 3.0 and 4.5 hours from the LKW (from 5.8% in 2008–2010 to 21.1% in 2017–2019) and in those with severe stroke (from 27.9% to 47.3%; Tables S3 and S4). However, it should be noted that the IVT rate remained low in patients arriving after 4.5 h from the LKW and in those with mild stroke (2.4% and 5.0% in 2017– 2019, respectively).

Over the 12 years, the MT rate almost tripled (from 3.2% to 9.2%), but the DPT did not improve. The increase in the MT rate was evident in patients arriving within 6 hours or between 6 and 12 hours of symptom onset and in those with moderate (NIHSS score 5–15) or severe stroke (Tables S3 and S4). However, it should be noted that the MT rates in patients arriving after 12 hours and in those with mild stroke remained low (2.1% and 1.9% in 2017– 2019, respectively).

Regarding secondary preventive treatments, the overall use of DAPT and DAPT in mild noncardioembolic stroke increased by 20.3% and 22.2%, respectively. The overall use of statins and the use of statins in LAA also increased by 19.7% and 19.2%, respectively. However, the overall use of anticoagulants and anticoagulants in atrial fibrillation decreased by 7.2% and 6.4%, respectively. Among types of anticoagulants in patients with atrial fibrillation, DOAC surpassed warfarin in the last epoch. Reimbursement for DOAC use in patients with atrial fibrillation was introduced by the Korean government in mid-2015. It should be mentioned that in ODEs, the use of anticoagulants decreased by 29.7%, and the use of DAPT increased by 20.3% (Table S5).

The clinical presentation and treatment trends did not change noticeably in subgroup analyses according to age group (not presented here) and sex (Table S6).

### Clinical Outcomes (Table 3)

We obtained information on clinical outcomes from 6,057 patients registered since 2011. The median follow-up duration was 367 days (interquartile range, 356-381 days). Out of these patients, 5861 (97.9%) had follow-up information post-discharge on time-to-event data, and 5809 (95.9%) had 3-month mRS scores. No significant difference was observed between patients with and without 3-month scores, except for a 1-point lower initial NIHSS median score, a higher prevalence of hypertension, and increased use of discharge statin among the former group (Table S8).

In-hospital death, the proportions of 3-month mRS scores 0–1 and mRS scores 0–2, and mortality did not exhibit significant trends before and after adjusting for age, sex, and initial NIHSS score. However, ordinal logistic regression revealed a significant increasing trend across the whole range of 3-month mRS scores after adjustments (common odds ratio, 1.18; 95% confidence interval, 1.04–1.33).

Further analyses were conducted to identify factors contributing to the lack of improvement in functional outcomes despite improvement in treatments. The proportion of mRS scores 0–1 increased in patients who underwent MT and those with severe strokes (from 33.7% to 47.8% and from 13.2% to 25.0%, respectively; Tables S9 and S10). However, the proportion of mRS scores 0–2 and the mortality did not demonstrate significant improvements in these subgroups.

The one-year cumulative incidence of recurrent stroke demonstrated a minor increase from 4.13% in 2011–2013 to 5.47% in 2017–2019 (p=0.04). However, this trend became non-significant in both a minimal model adjusting for age and sex and in the model adjusting for sex, age, stroke risk factors, and stroke subtypes. As post-hoc analysis, pairwise comparisons were made between the three epochs. Notably, the increase in stroke recurrence between 2014-16 and 2017-19 remained significant after Bonferroni adjustment (p=0.01). However, the increase between 2011-13 and 2014-16, or 2011-13 and 2017-19 was no longer significant (p=0.90 and p-0.12, respectively).

Figure 2 demonstrates the incidence of recurrent stroke according to the index stroke subtypes. The increase in the stroke recurrence rate was observed in CES, ODE and UDE. In subgroup analysis of ischemic stroke subtype, the cumulative incidence of recurrent stroke increased significantly only in CES and UDE (p_trend_ =0.02 and p_trend_ =0.03, respectively, Table S11). Notably, in the CES group, patients who took warfarin had a lower recurrence rate than those who did not (Figure S1, p=0.14 on a log-rank test). In the ODE group, there was no difference in stroke recurrence between patients who took warfarin and those who did not (Figure S2).

**Figure 2.**
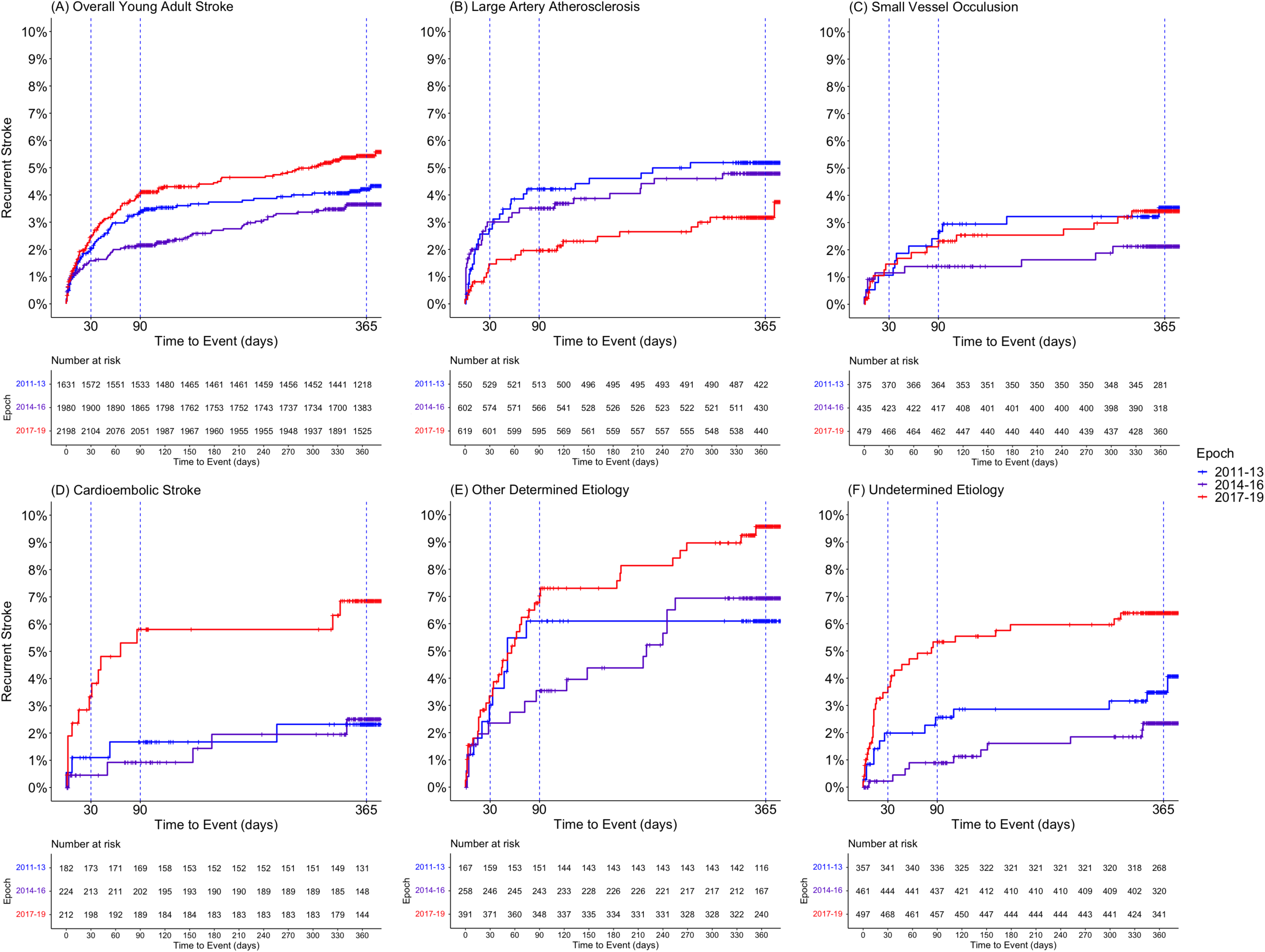
Stroke Recurrence According to Epochs and Stroke Subtypes Stroke recurrence in (A) overall, (B) large artery atherosclerosis, (C) small vessel occlusion, (D) cardioembolic stroke, (E) other determined etiology, and (F) undetermined etiology.

## Discussion

In our nationwide cohort of young adults with AIS, parameters regarding advanced treatments such as IVT and MT rates, DNT, and secondary preventive treatments, including DAPT for mild noncardioembolic stroke, DOAC for atrial fibrillation, and statin for LAA, improved over the 12 years. However, the 3-month mRS score, 1-year mortality, and 1-year stroke recurrence rate did not improve. The stroke recurrence rate even increased in the last epoch, which was attributed to increased recurrence in patients with CES, ODE, or UDE.

In our study, the functional outcomes at 3 months post-AIS in young adults did not improve and even showed signs of worsening after adjusting for age, sex, and initial stroke severity. The proportion of mRS scores 0–2 was 84% in our study, which aligns with the percentages reported from the late 1990s to mid 2010s in studies of young patients with ischemic stroke (84–94%).^13, 14, 16^ Interestingly, some countries have observed improvement in functional outcomes over the last two decades in the ischemic stroke population across all ages.^11, 21–23^ Increased proportions of 3-month mRS scores 0–2 were reported in China (from 37% in 2002 to 71% in 2016) and Spain (from 49% in 2008 to 55% in 2016).^21, 22^ The Riks-Stroke registry and Get With The Guideline Stroke data also demonstrated improving trend in discharge to home as a surrogate measure of functional outcome.^11, 23^

The unchanged trend in functional outcomes despite the enhanced provision of treatments may be attributed to a ceiling effect; the majority of young patients were already functionally independent in the early epoch (3-month mRS scores 0–2, 83.5% in 2011–2013). On the other hand, the IVT rate and DNT have not shown significant improvement between 2011-2013 and 2017-2019 (13.3% vs. 13.8% and 40 min vs. 39 min, respectively). This indicates that patients who could potentially benefit from IVT may not receive optimal treatment, as the hospital arrival time from the onset of stroke (ODT) has remained relatively stagnant. In addition, even though the MT rate has doubled, the number of patients benefiting from this therapy is still limited, accounting for less than 10% of AIS patients even in 2017-19.

Thus, more emphasis should be placed on areas that would benefit a larger population, such as enhancing awareness and treatment of risk factors. This may have a more substantial impact on improving outcomes than merely extending the reach of reperfusion therapy. It is worth noting that only half of our study subjects with known hypertension or diabetes were received medication before their index stroke. Recent studies have underscored that vascular risk factors in young adults are often undertreated, partly because the current treatment thresholds for these risk factors may underestimate the actual risk of stroke in this younger population.^2^

We demonstrated that the stroke recurrence rate among young patients did not decrease and was even highest in the last epoch. This finding contrasts the decreasing trend observed among young adults with AIS in Sweden^16^ and the AIS population across all ages in the United States before the early 2010s.^25^ The apparent increase in stroke recurrence in the last epoch might be influenced by a low number of events and potential changes in the study population characteristics over time. Further studies in other populations are needed to ascertain if this trend may be replicated.

In an attempt to discern potential factors contributing to the highest stroke recurrence rate in the most recent epoch, we scrutinized changes in risk factors, stroke subtypes, and preventive treatments across all epochs. Over the 12 years, we identified three significant changes: a doubling in the prevalence of obesity, an increase in ODE among ischemic stroke subtypes, and a decrease in the use of warfarin (Tables 1, 2, and Table S5). The increase in obesity in young adults is consistent with trends seen in several developed countries, including the United States and South Korea.^25–27^ However, it is worth noting that the increase in the prevalence of obesity in young stroke patients was slightly steeper than that found in the general population of young adults according to data from the Korean National Health Insurance Service (from 4.0% to 8.5% between 2009 and 2019).^25^

**Table 1.**
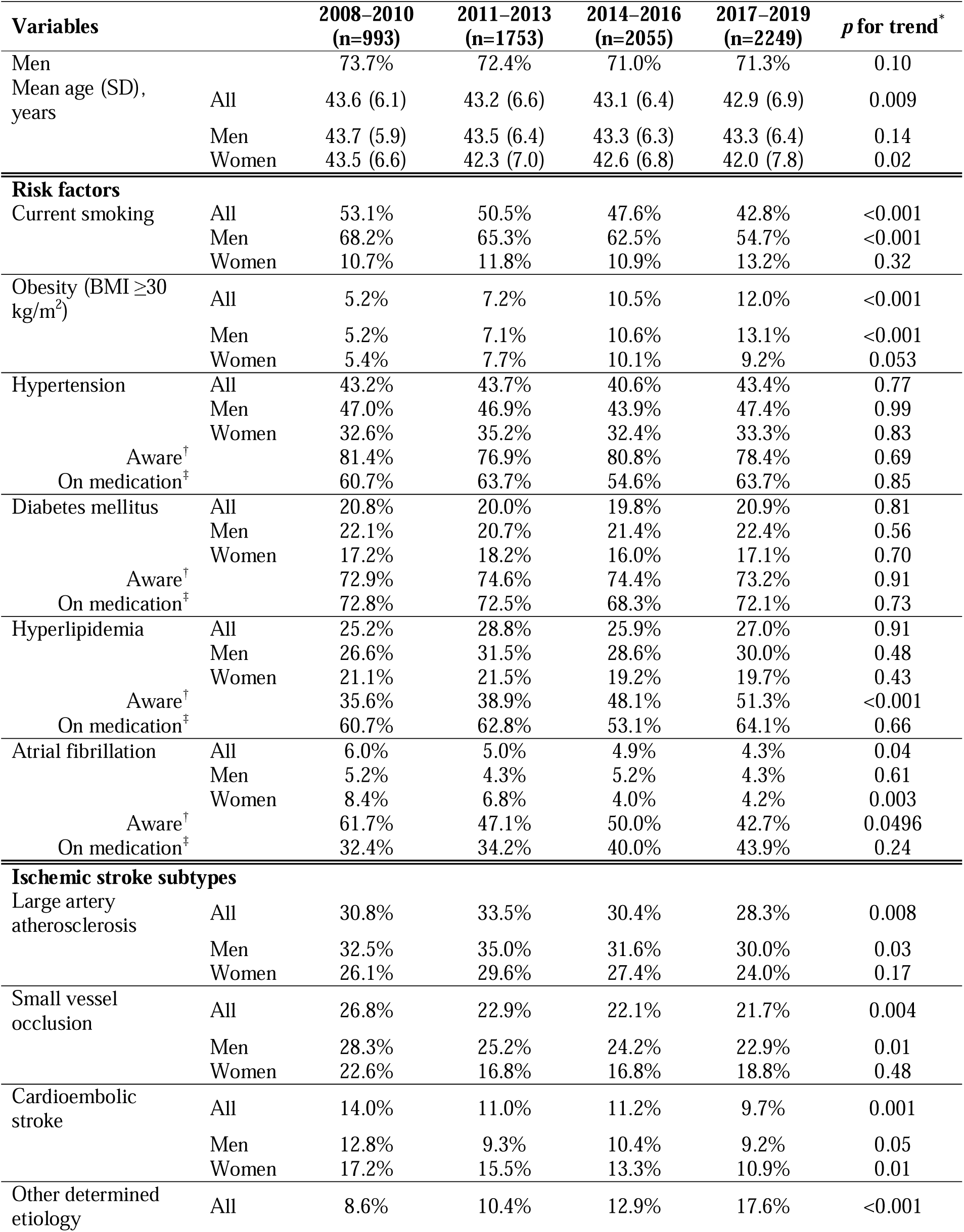
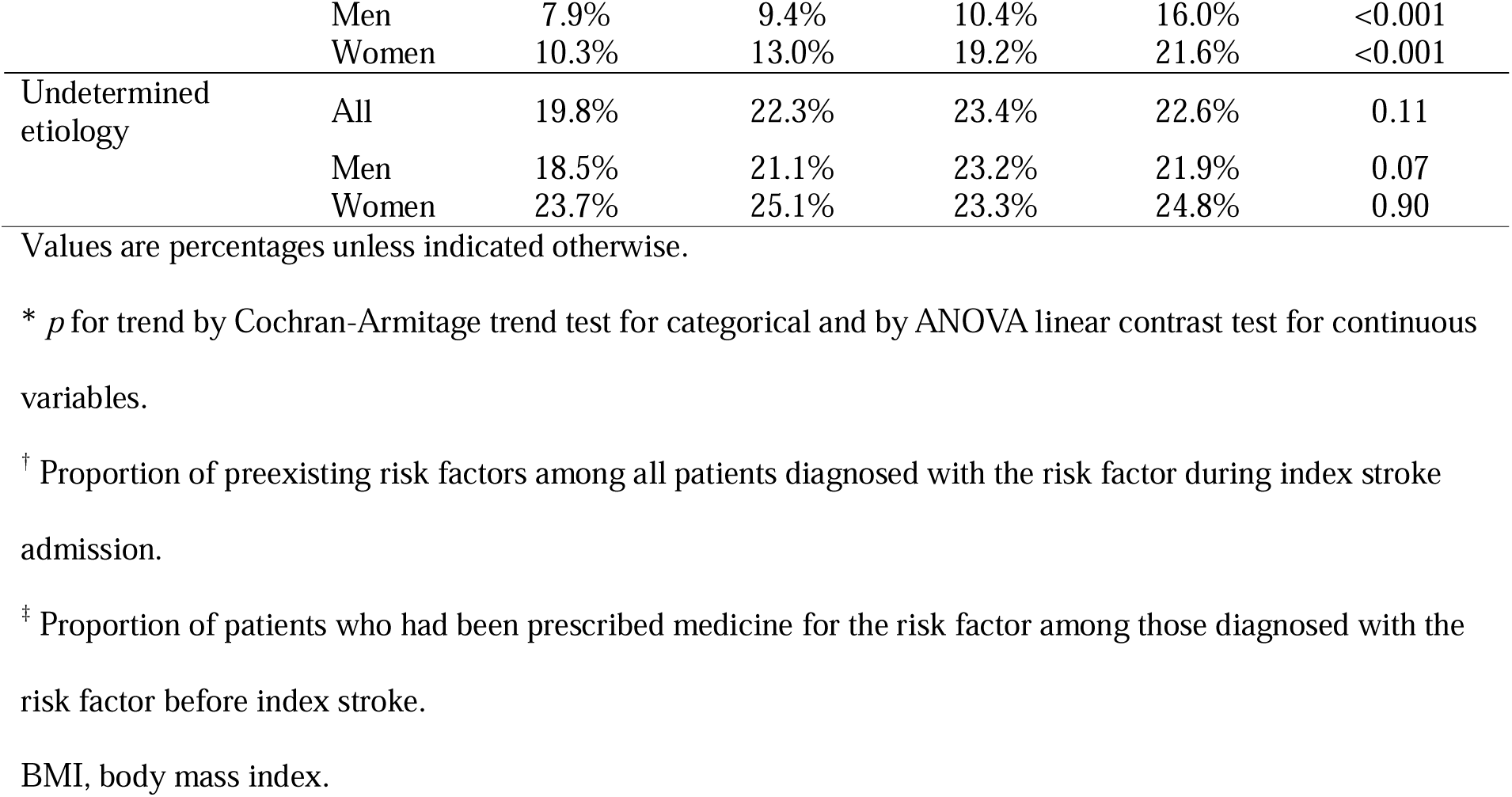
Secular Trends in Demographics, Risk Factors, and Subtypes of Ischemic Stroke.

**Table 2.** Secular Trends in Clinical Presentation and Treatments.

| Variables | 2008–2010<br>(n=993) | 2011–2013<br>(n=1753) | 2014–2016<br>(n=2055) | 2017–2019<br>(n=2249) | *p* for trend* |
| --- | --- | --- | --- | --- | --- |
| **Onset-to-door time** |  |  |  |  |  |
| Median FAT-to-door (IQR), h | 8.4 (1.9–29.5) | 8.2 (2.4–33.5) | 9.4 (2.1–33.6) | 8.0 (2.0–31.6) | 0.66 |
| Median LKW-to-door (IQR), h | 11.7 (2.8–32.7) | 11.1 (3.0–37.1) | 12.5 (3.1–38.7) | 11.4 (2.8–34.6) | 0.62 |
| LKW-to-door within 3.5h | 27.7% | 27.8% | 26.5% | 28.1% | 0.92 |
| **Stroke severity** |  |  |  |  |  |
| Mean initial NIHSS score (SD) | 4.8±5.5 | 4.2±5.3 | 4.2±5.1 | 4.0±4.9 | <0.001 |
| Median initial NIHSS score (IQR) | 3 (1–6) | 2 (1–5) | 2 (1–5) | 2 (1–5) | <0.001 |
| Mild stroke (≤4) | 64.9% | 71.0% | 70.7% | 69.8% | 0.06 |
| Moderate stroke (5–15) | 28.3% | 23.4% | 24.0% | 25.3% | 0.39 |
| Severe stroke (16–42) | 6.8% | 5.6% | 5.4% | 4.9% | 0.03 |
| **Reperfusion therapy** |  |  |  |  |  |
| IVT rate | 9.5% | 13.3% | 14.0% | 13.8% | 0.004 |
| Median DNT (IQR), min | 58 (43–83) | 40 (31–54) | 37 (26.8–49) | 39 (29–55.8) | <0.001 |
| DNT ≤60 min | 56.2% | 84.2% | 86.3% | 81.5% | <0.001 |
| MT rate | 3.2% | 5.3% | 7.5% | 9.2% | <0.001 |
| Median DPT (IQR), min | 113 (88–146) | 110 (92–137) | 115.5 (87.3–158.5) | 106 (85–153.3) | 0.84 |
| DPT ≤90 min | 26.1% | 23.5% | 31.2% | 30.9% | 0.28 |
| **Discharge medications** |  |  |  |  |  |
| Dual antiplatelet therapy | 26.7% | 24.8% | 32.8% | 47.0% | <0.001 |
| In noncardioembolic minor stroke | 28.6% | 25.0% | 35.2% | 50.8% | <0.001 |
| Anticoagulation | 15.8% | 13.1% | 9.6% | 8.6% | <0.001 |
| Warfarin | 15.2% | 11.6% | 6.8% | 4.3% | <0.001 |
| DOAC | 0.0% | 0.2% | 2.3% | 3.8% | <0.001 |
| Anticoagulation in AF | 78.3% | 78.2% | 71.0% | 71.9% | 0.22 |
| Warfarin in AF | 75.0% | 72.4% | 41.0% | 14.6% | <0.001 |
| DOAC in AF | 0.0% | 4.6% | 30.0% | 56.2% | <0.001 |
| Statin | 71.5% | 79.9% | 86.9% | 91.2% | <0.001 |
| Among LAA | 76.1% | 86.2% | 89.8% | 95.3% | <0.001 |

Values are percentages unless indicated otherwise.
\* *p* for trend by Cochran–Armitage trend test for categorical and ANOVA linear contrast test for continuous
variables.
AF, atrial fibrillation; DNT, door-to-needle time; DOAC, direct oral anticoagulant; DPT, door-to-puncture time;
FAT, first abnormal time; IVT, intravenous thrombolysis; LAA, large artery atherosclerosis; LKW, last known
well time; MT, mechanical thrombectomy; National Institute of Health Stroke Scale.

**Table 3.**
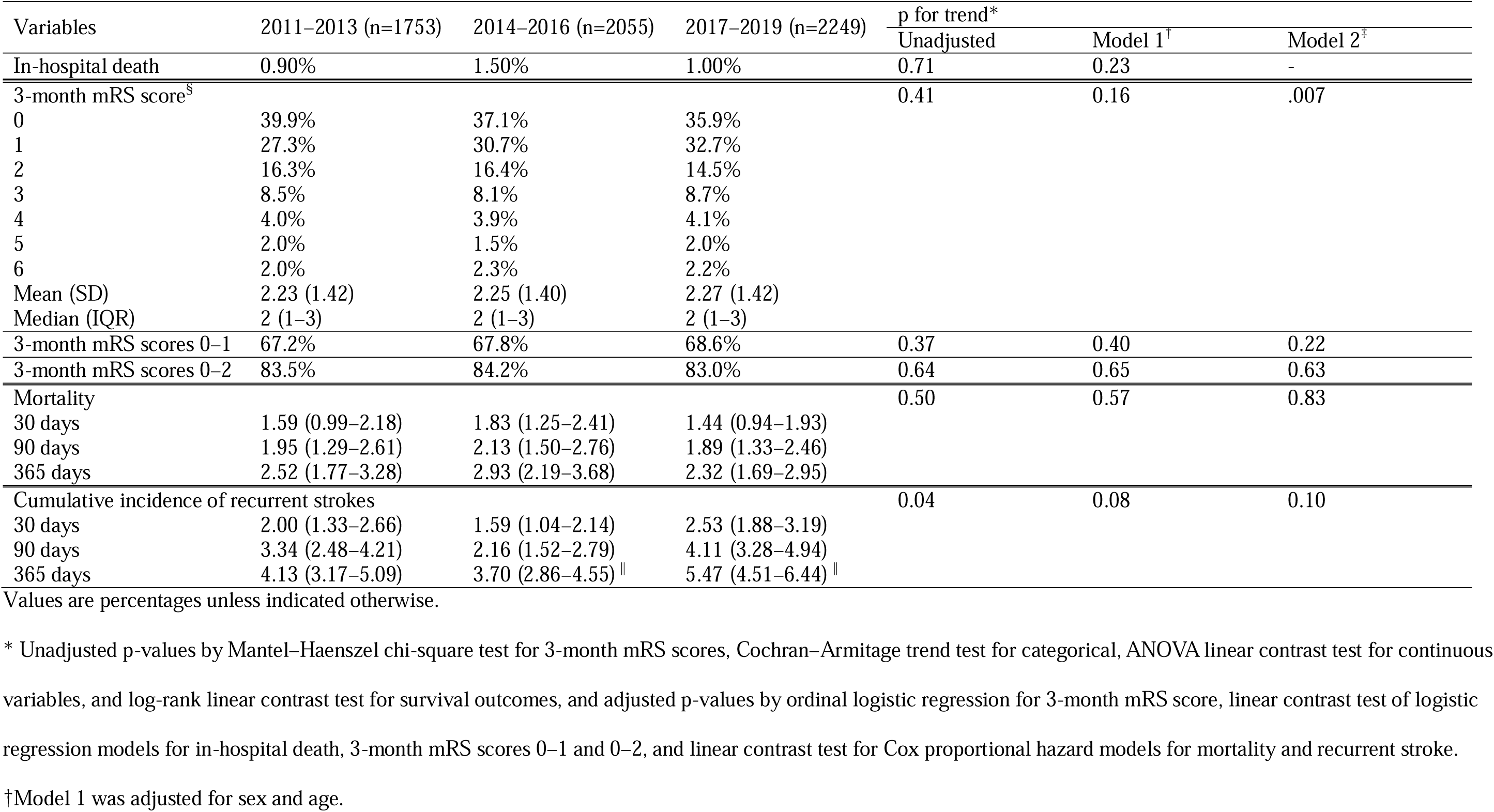
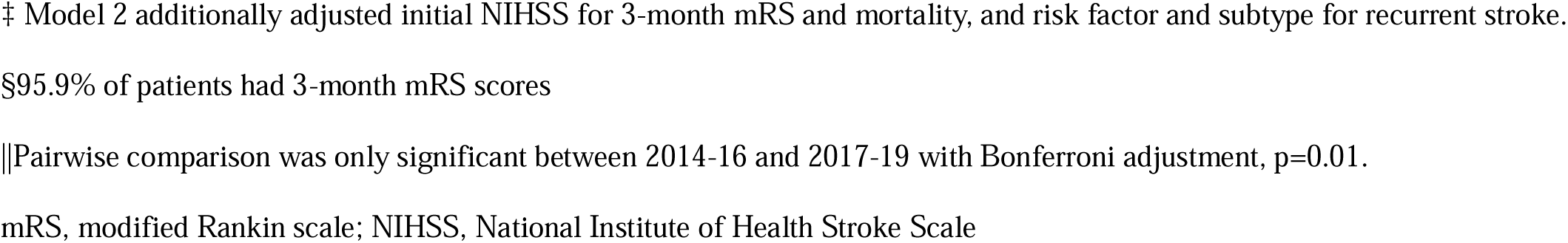
Secular Trends in Clinical Outcomes of Ischemic Stroke.

| Variables | 2011–2013 (n=1753) | 2014–2016 (n=2055) | 2017–2019 (n=2249) | p for trend* |  |  |
| --- | --- | --- | --- | --- | --- | --- |
|  |  |  |  | Unadjusted | Model 1<sup>†</sup> | Model 2<sup>‡</sup> |
| In-hospital death | 0.90% | 1.50% | 1.00% | 0.71 | 0.23 | - |
| 3-month mRS score<sup>§</sup> |  |  |  | 0.41 | 0.16 | .007 |
| 0 | 39.9% | 37.1% | 35.9% |  |  |  |
| 1 | 27.3% | 30.7% | 32.7% |  |  |  |
| 2 | 16.3% | 16.4% | 14.5% |  |  |  |
| 3 | 8.5% | 8.1% | 8.7% |  |  |  |
| 4 | 4.0% | 3.9% | 4.1% |  |  |  |
| 5 | 2.0% | 1.5% | 2.0% |  |  |  |
| 6 | 2.0% | 2.3% | 2.2% |  |  |  |
| Mean (SD) | 2.23 (1.42) | 2.25 (1.40) | 2.27 (1.42) |  |  |  |
| Median (IQR) | 2 (1–3) | 2 (1–3) | 2 (1–3) |  |  |  |
| 3-month mRS scores 0–1 | 67.2% | 67.8% | 68.6% | 0.37 | 0.40 | 0.22 |
| 3-month mRS scores 0–2 | 83.5% | 84.2% | 83.0% | 0.64 | 0.65 | 0.63 |
| Mortality |  |  |  | 0.50 | 0.57 | 0.83 |
| 30 days | 1.59 (0.99–2.18) | 1.83 (1.25–2.41) | 1.44 (0.94–1.93) |  |  |  |
| 90 days | 1.95 (1.29–2.61) | 2.13 (1.50–2.76) | 1.89 (1.33–2.46) |  |  |  |
| 365 days | 2.52 (1.77–3.28) | 2.93 (2.19–3.68) | 2.32 (1.69–2.95) |  |  |  |
| Cumulative incidence of recurrent strokes |  |  |  | 0.04 | 0.08 | 0.10 |
| 30 days | 2.00 (1.33–2.66) | 1.59 (1.04–2.14) | 2.53 (1.88–3.19) |  |  |  |
| 90 days | 3.34 (2.48–4.21) | 2.16 (1.52–2.79) | 4.11 (3.28–4.94) |  |  |  |
| 365 days | 4.13 (3.17–5.09) | 3.70 (2.86–4.55) | 5.47 (4.51–6.44) |  |  |  |

Values are percentages unless indicated otherwise.
\* Unadjusted p-values by Mantel–Haenszel chi-square test for 3-month mRS scores, Cochran–Armitage trend test for categorical, ANOVA linear contrast test for continuous variables, and log-rank linear contrast test for survival outcomes, and adjusted p-values by ordinal logistic regression for 3-month mRS score, linear contrast test of logistic regression models for in-hospital death, 3-month mRS scores 0–1 and 0–2, and linear contrast test for Cox proportional hazard models for mortality and recurrent stroke.
<sup>†</sup>Model 1 was adjusted for sex and age.
‡ Model 2 additionally adjusted initial NIHSS for 3-month mRS and mortality, and risk factor and subtype for recurrent stroke.
§95.9% of patients had 3-month mRS scores
||Pairwise comparison was only significant between 2014-16 and 2017-19 with Bonferroni adjustment, $p=0.01$ .
mRS, modified Rankin scale; NIHSS, National Institute of Health Stroke Scale

The changes in the usage patterns of antithrombotic therapies might have played a role in the stagnant trend in stroke recurrence rates among young patients. We observed a decline in the use of warfarin among patients with CES, ODE, and UDE, with a reduction of - 24.3%, -17.8%, and -7.3%, respectively, between 2011-13 and 2017-19. Interestingly, these are the same three subtypes that exhibited increased stroke recurrence in the most recent epoch (Figure 2, Table S5, and Table S11). Additionally, a lower recurrence rate was found in CES patients on warfarin than those on other antithrombotic therapies (Figure S1). Guidelines specific to young patients with CES and no atrial fibrillation, ODE, or UDE are not well-established.^8^ In some patients with congenital or valvular heart disease, warfarin may be superior to antiplatelets or DOACs by blocking several coagulation pathways simultaneously.^28^ Further research is warranted to verify the recent increase in stroke recurrence in young adults and the influence of warfarin use.

Over the 12 years, the mean age of young patients with AIS decreased, which was attributed to an increase in the proportion of the youngest group (women, 18–30 years) (Figure 1B). A recent systematic review reported that the risk of stroke was higher in young women (15–35 years) than in young men.^29^ However, despite the increasing trend in the proportion of women in our study, there were more men. A preponderance of young male patients with stroke has been reported in other studies from Asian countries.^30^

This study has several limitations. Firstly, most participating hospitals in our registry are either university hospitals or regional stroke centers, which may not necessarily represent the entirety of acute stroke care hospitals in South Korea. However, it is noteworthy that the demographics and baseline characteristics in our study do not significantly deviate from a previous study that relied on national claims data,^31^ which includes 98–99% of the total Korean population. Secondly, there was an increase in the number of hospitals participating in our registry throughout the study period. This could have potential implications for secular trend analysis and should be considered when interpreting our results as certain factors (e.g., socioeconomic status) could change over time. Thirdly, given the retrospective nature of our data collection and the descriptive study design, we could not establish causal relationship between treatments and outcomes. Fourthly, we could not determine the causal relationship between treatments and outcomes due to the retrospective nature of data collection and the descriptive study design. Fifthly, since we started to collect outcome data in 2011, we could only analyze the trends in outcomes during the last three epochs. Our findings of an increase in stroke recurrence in the last epoch could be due to chance; however, this finding may warrant further studies with a more extended observation period. Lastly, our study was based on a nationwide stroke registry in South Korea. Therefore, the generalizability of our findings to other countries and ethnicities is limited.

In summary, clinical outcomes of young adults with AIS in South Korea did not improve between 2011 and 2019 despite increased delivery of AIS and recurrent preventive treatments. Potential areas of improvement to close the clinical gap include early detection and treatment of traditional risk factors, expanded implementation of reperfusion therapy, reduction in delays in the administration of reperfusion therapy, and establishment of optimal antithrombotic strategies in young patients with CES, ODE, and UDE.

## Data Availability

The data used in this study may be made available upon request to the corresponding author.

