## Supplemental material for "Improvement in Delivery of Ischemic Stroke Treatments but Stagnation of Clinical Outcomes in Young Adults in South Korea"

**Table S1.** Selected Variables, Their Definition, and Missing Counts.

**Table S2**. Proportions of Other Determined Etiology Subtypes.

**Table S3**. IV Thrombolysis or Mechanical Thrombectomy Rate According to Initial Stroke Severity.

**Table S4**. IV Thrombolysis or Mechanical Thrombectomy Rate According to Time from Last Normal to Hospital Arrival.

**Table S5** Secular Trends in Discharge Antithrombotics According to Ischemic Stroke Subtypes

**Table S6**. Clinical Presentation and Treatments in Young Adults with Ischemic Stroke Stratified by Sex

**Table S7**. Clinical Outcomes Stratified by Sex.

**Table S8**. Comparison between patients with and without 3 month mRS.

**Table S9**. 3-months Functional Outcome and Case-fatality After Stroke According to a Mode of Reperfusion Therapy.

**Table S10**. 3-months Functional Outcome and Case-fatality After Stroke According to Initial Stroke Severity.

**Table S11**. Subgroup analysis of stroke recurrence stratified by ischemic stroke subtype.

**Figure S1**. Stroke Recurrence In Cardioembolic Stroke According to Discharge Medication (Warfarin vs. Others).

**Figure S2**. Stroke Recurrence In Other Determined Etiologies According to Discharge Medication (Warfarin vs. Others).

**Table S1.** Selected Variables, Their Definition, and Missing Counts.

| **Variables** | **Definition** | **Missing** |
|---|---|---|
| Sex | The biological sex of patient (men or women) | 0 |
| Age | The age of patient at the timeof index stroke admission | 0 |
| Admission year | The calendar year of the patient's admission | 0 |
| First abnormal to arrival time | The time interval (in minutes) from the first recognized symptoms of stroke to the patient's arrival at the hospital. | 0 |
| Last normal to arrival time | The time interval (in minutes) from the patient's last identified baseline state before the index stroke to their arrival at the hospital. | 0 |
| Prior disability | The patient's functional state before the index stroke as measured by the modified Rankin scale (mRS) | 145 |
| Initial NIHSS | The initial stroke severity as measured by the NIH Stroke Scale (NIHSS) | 0 |
| History of past TIA or stroke | A history of any previous transient ischemic attacks (TIAs) or strokes | 0 |
| History of hypertension | The patient's hypertension status based on following criteria: 1) patients who was prescribed antihypertensive before index stroke, 2) patients who had been diagnosed hypertension before index stroke, 3) when patient's blood pressure was 140/90 or higher after acute period, requiring long-term antihypertensive. | 0 |
| Aware hypertension | The number of patients diagnosed with hypertension before index stroke admission | 0 |
| 'On medication' hypertension | The number of patients who treated with antihypertensive medication before index stroke admission | 0 |
| History of diabetes | The patient's diabetes status based on following criteria: 1) patients who was prescribed antidiabetic agent before index stroke, 2) patients who had been diagnosed diabetes before index stroke, 3) when patient's fasting glucose level was 126 mg/dL or higher, or postprandial glucose level 200 mg/dL or higher after acute phase, or when patient's glucose level 200 mg/dL with diabetic symptoms. | 0 |
| Aware diabetes | The number of patients diagnosed with diabetes before index stroke admission | 0 |
| 'On medication' diabetes | The number of patients who treated with diabetes medication before index stroke admission | 0 |
| History of hyperlipidemia | The patient's hyperlipidemia status based on following criteria: 1) patients who was prescribed statin or other hyperlipidemia medicine before index stroke, 2) patients who had been diagnosed hyperlipidemia before index stroke, 3) when patient's fasting LDL cholesterol level was 160 mg/dL or higher, or total cholesterol level 240 mg/dL or higher after acute phase. | 0 |
| Aware hyperlipidemia | The number of patients diagnosed with hyperlipidemia before index stroke admission | 0 |
| 'On medication' hyperlipidemia | The number of patients who treated with hyperlipidemia medication before index stroke admission | 0 |
| History of atrial fibrillation | A history of atrial fibrillation, including both persistent and paroxysmal forms | 0 |
| Aware atrial fibrillation | The number of patients diagnosed with atrial fibrillation before index stroke admission | 0 |
| 'On medication' atrial fibrillation | The number of patients who treated with anticoagulants before index stroke admission | 0 |
| Current smoking | The patient's current smoking status (not including ex-smokers) | 1 |
| Obesity | Obesity was defined as a calculated BMI of 30 kg/m2 or higher | 129 |
| Systolic blood pressure /diastolic blood pressure | The patient's blood pressure measured in mmHg at the initial days of hospitalization | 27 |

| | | |
|---|---|---|
| HDL cholesterol | HDL cholesterol level measured at the initial days of hospitalization (in mg/dL) | 251 |
| LDL cholesterol | LDL cholesterol level measured at the initial days of hospitalization (in mg/dL) | 251 |
| Triglyceride | Triglyceride level measured at the initial days of hospitalization (in mg/dL) | 213 |
| Random glucose | Blood glucose level measured at the initial days of hospitalization (in mg/dL) | 80 |
| Length of hospitalization | The lengh of hospital stay from admission to discharge (in days) | 2 |
| TOAST classification | The stroke etiology as classified by the Trial of Org 10172 in Acute Stroke Treatment (TOAST):<br>Large artery occlusion, small vessel occlusion, cardioembolic stroke, other determined etiology, undetermined etiology (two or more, negative result on evaluation, incomplete evaluation) | 1 |
| Detailed diagnosis of other determined etiology | Since MRI-based TOAST classification algorithm was introduced in our registry in July 2011, detailed diagnoses of other determined etiology stroke were captured as following categories:<br>1) Arterial dissection, 2) infectious or inflammatory disorder, 3) intrinsic vascular disorder such as Moyamoya disease, 4) hemostatic disorder, 5) migraine-related stroke, 6) hereditary disease such as CASASIL, 7) hypoperfusional stroke, 8) iatrogenic, and 9) others. | 69/814 |
| MRI | The rate of MRI conduction during hospitalization | 0 |
| CTA or MRA | The rate of either CT or MR angiography conduction during hospitalization | 0 |
| Transthoracic echocardiography | The rate of TTE conduction during hospitalization | 0 |
| Transesophageal echocardiography | The rate of TEE conduction during hospitalization | 0 |
| Holter monitor | The rate of Holter monitor conduction during hospitalization | 0 |
| Mode of perfusion therapy | The type of acute reperfusion therapy, including none, intravenous (IV) thrombolysis only, mechanical thrombectomy only, or both IV thrombolysis and mechanical thrombectomy | 0 |
| Arrival to IV thrombolysis time | The time interval (in minutes) from patient arrival at the hospital to the start of IV thrombolysis for patients who received this therapy in participating hospitals. | 3/927 |
| Arrival to mechanical thrombectomy time | The time interval (in minutes) from patient arrival at the hospital to the start of mechanical thrombectomy for patients who received this therapy in participating hospitals | 7/487 |
| Discharge antiplatelet use | The type and number of antiplatelet drugs prescribed at discharge, including aspirin, clopidogrel, aspirin + dipyridamole, cilostazol, triflusal, ticlopidine, or others | 0 |
| Discharge anticoagulant use | The type and number of anticoagulants prescribed at discharge, including warfarin, apixaban, dabigatran, rivaroxaban, edoxaban, or others | 0 |
| Discharge statin use | Whether a statin was prescribed at discharge or not | 4 |
| In-hospital death | The number of all-cause deaths that occurred during hospitalization for the index stroke | 0 |
| 3-month functional outcome | The patient's functional status as measured by the modified Rankin Scale (mRS) collected 3 months after the index stroke. The mRS was used in the analysis as an ordinal variable or dichotomized into 0-1 and 0-2. | 248/6057 |
| Recurrent stroke event | The occurrence of first recurrent stroke, including ischemic stroke, hemorrhagic stroke, or transient ischemic attack (TIA), was captured at discharge, 3 months, and 1 year after the index | 0/6057 |

| | | |
|---|---|---|
| | stroke. | |
| Time to stroke recurrence | The time interval (in days) from the index stroke to the recurrent stroke | 0/6057 |
| All-cause death | The occurence of deaths, including both vascular and nonvascular events, was captured at discharge, 3 months, and 1 year after the index stroke. | 0/6057 |
| Time to all-cause death | The time interval (in days) from the index stroke to all-cause death | 0/6057 |

**Table S2**. Presumed Causes of Stroke in Patients with Other Determined Etiology*

| | Sex | 2011-2013 (n=1116) | 2014-2016 (n=1904) | 2017-2019 (n=2102) | P for trend† |
|---|---|---|---|---|---|
| Arterial dissection | All | 4.9% | 8.3% | 9.8% | < .001 |
| | Men | 5.7% | 7.9% | 10.4% | < .001 |
| | Women | 3.1% | 9.2% | 8.2% | 0.02 |
| Intrinsic disease of arterial wall | All | 2.6% | 2.7% | 4.5% | .001 |
| | Men | 1.4% | 1.4% | 3.1% | .002 |
| | Women | 5.6% | 5.9% | 7.9% | 0.14 |
| Hemostatic disease/coagulopathy | All | 0.2% | 0.4% | 1.2% | <.001 |
| | Men | 0.1% | 0.4% | 0.7% | 0.03 |
| | Women | 0.3% | 0.5% | 2.3% | .003 |
| Cancer-related coagulopathy | All | 0% | 0.4% | 1.0% | <.001 |
| | Men | 0% | 0.4% | 0.9% | .005 |
| | Women | 0% | 0.4% | 1.2% | 0.02 |
| Cerebral venous thrombosis | All | 0.6% | 0.2% | 0.8% | 0.34 |
| | Men | 0.4% | 0.1% | 0.1% | 0.52 |
| | Women | 1.2% | 0.4% | 1.5% | 0.48 |
| Genetic/Hereditary disease | All | 0.5% | 0.6% | 0.7% | 0.67 |
| | Men | 0.6% | 0.7% | 0.5% | 0.73 |
| | Women | 0.3% | 0.5% | 1.0% | 0.20 |
| Infectious disease | All | 0.4% | 0.3% | 0.4% | 0.69 |
| | Men | 0.3% | 0.1% | 0.3% | 0.58 |
| | Women | 0.6% | 0.7% | 0.7% | 0.98 |
| Iatrogenic | All | 1.0% | 0.2% | 0.2% | .004 |
| | Men | 0.8% | 0.2% | 0.2% | .046 |
| | Women | 1.6% | 0.2% | 0.3% | 0.04 |
| Migraine-related | All | 0.2% | 0.1% | 0.1% | 0.93 |
| | Men | 0% | 0.1% | 0.1% | 0.29 |
| | Women | 0.6% | 0% | 0.2% | 0.23 |
| Hypoperfusion syndrome | All | 0.5% | 0.2% | 0% | <.001 |
| | Men | 0% | 0% | 0% | - |
| | Women | 1.9% | 0.5% | 0% | <.001 |

* Among patients who were investigated specific diagnosis by Magnetic resonance imaging-based diagnostic AlGorithm for acute Ischemic stroke subtype Classification (MAGIC) protocol.

† P for trend by Cochran-Armitage trend test

**Table S3**. IV Thrombolysis or Mechanical Thrombectomy Rate According to Time from Last Normal to Hospital Arrival.

| **Reperfusion therapy** | ***Last known normal-*** **to-door time** | **2008-2010 (n=993)** | **2011-2013 (n=1753)** | **2014-2016 (n=2055)** | **2017-2019 (n=2249)** | **P for trend*** |
|---|---|---|---|---|---|---|
| IVT | ≤ 3 h | 30.8% | 42.4% | 43.6% | 42.1% | 0.02 |
| | 3 ~ 4.5 h | 5.8% | 19.7% | 22.1% | 21.1% | 0.06 |
| | > 4.5 h | 1.6% | 2.0% | 2.8% | 2.4% | 0.18 |
| MT | ≤ 6 h | 6.4% | 11.4% | 16.8% | 18.7% | <.001 |
| | 6 ~ 12 h | 2.8% | 2.8% | 6.0% | 9.5% | <.001 |
| | > 12 h | 1.0% | 1.3% | 1.4% | 2.1% | 0.08 |

* P-value by Cochran-Armitage trend test

IVT, intravenous thrombolysis; MT, mechanical thrombectomy.

**Table S4**. IV Thrombolysis or Mechanical Thrombectomy Rate According to Baseline Stroke Severity.

| Reperfusion therapy | Initial stroke severity | 2008-2010 (n=993) | 2011-2013 (n=1753) | 2014-2016 (n=2055) | 2017-2019 (n=2249) | P for trend* |
|---|---|---|---|---|---|---|
| IVT | Mild (0-4) | 2.3% | 4.5% | 5.2% | 5.0% | 0.02 |
| | Moderate (5-15) | 21.4% | 34.4% | 34.5% | 31.8% | 0.02 |
| | Severe (16-42) | 27.9% | 37.8% | 38.2% | 47.3% | 0.01 |
| MT | Mild (0-4) | 0.3% | 1.2% | 1.7% | 1.9% | 0.01 |
| | Moderate (5-15) | 5.7% | 11.0% | 16.6% | 21.1% | <.001 |
| | Severe (16-42) | 20.6% | 33.7% | 43.6% | 53.6% | <.001 |

* P-value by Cochran-Armitage trend test
IVT, intravenous thrombolysis; MT, mechanical thrombectomy.

**Table S5** Secular Trends in Discharge Antithrombotics According to Ischemic Stroke Subtypes

| Stroke subtype | Discharge prescription | 2008–2010 | 2011–2013 | 2014–2016 | 2017–2019 | *p* for trend* |
|---|---|---|---|---|---|---|
| Large artery atherosclerosis (n=2155) | Antiplatelet | 88.6% | 93.2% | 95.0% | 97.2% | <0.001 |
| | Aspirin + clopidogrel | 38.6% | 36.0% | 46.6% | 61.9% | <0.001 |
| | Anticoagulant | 7.8% | 4.9% | 1.9% | 1.3% | <0.001 |
| | Warfarin | 7.5% | 3.6% | 1.8% | 0.6% | <0.001 |
| | DOAC | 0% | 0% | 0% | 0.5% | 0.03 |
| Small vessel occlusion (n=1609) | Antiplatelet | 97.4% | 96.0% | 96.9% | 98.7% | 0.08 |
| | Aspirin + clopidogrel | 22.9% | 17.5% | 30.2% | 49.0% | <0.001 |
| | Anticoagulant | 2.3% | 2.5% | 0.2% | 0.8% | 0.03 |
| | Warfarin | 1.9% | 1.8% | 0% | 0.6% | 0.02 |
| | DOAC | 0% | 0% | 0.2% | 0.2% | 0.30 |
| Cardioembolic stroke (n=781) | Antiplatelet | 38.1% | 56.5% | 51.1% | 49.1% | 0.24 |
| | Aspirin + clopidogrel | 11.5% | 10.4% | 12.6% | 16.1% | 0.12 |
| | Anticoagulant | 55.4% | 49.2% | 46.8% | 49.5% | 0.30 |
| | Warfarin | 54.0% | 47.7% | 32.9% | 23.4% | <0.001 |
| | DOAC | 0% | 1.0% | 13.0% | 23.9% | <0.001 |
| Other determined etiology (n=929) | Antiplatelet | 56.7% | 76.4% | 83.8% | 86.4% | <0.001 |
| | Aspirin + clopidogrel | 22.4% | 25.3% | 35.3% | 42.7% | <0.001 |
| | Anticoagulant | 38.8% | 25.3% | 13.5% | 9.1% | <0.001 |
| | Warfarin | 38.8% | 23.6% | 11.3% | 5.8% | <0.001 |
| | DOAC | 0% | 0% | 0.8% | 1.8% | 0.26 |
| Undetermined etiology (n=1576) | Antiplatelet | 86.6% | 83.9% | 85.6% | 87.3% | 0.43 |
| | Aspirin + clopidogrel | 25.9% | 22.3% | 25.5% | 43.3% | <0.001 |
| | Anticoagulant | 8.6% | 12.6% | 8.6% | 7.5% | 0.09 |
| | Warfarin | 7.6% | 10.5% | 4.8% | 2.9% | <0.001 |
| | DOAC | 0% | 0.5% | 2.9% | 4.3% | <0.001 |

* *p*-value by Cochran–Armitage trend test.
DOAC, direct oral anticoagulants

**Table S6**. Clinical Presentation and Treatments in Young Adults with Ischemic Stroke Stratified by Sex.

| **Variables** | | **2008-2010 (n=993)** | **2011-2013 (n=1753)** | **2014-2016 (n=2055)** | **2017-2019 (n=2249)** | **p for trend*** |
|---|---|---|---|---|---|---|
| **LKW-to-door within 3.5 h** | All | 27.7% | 27.8% | 26.5% | 28.1% | 0.92 |
| | Men | 25.4% | 26.6% | 26.0% | 28.2% | 0.17 |
| | Women | 34.1% | 31.1% | 27.7% | 27.8% | **0.04** |
| **Stroke severity** | | | | | | |
| Mild stroke (NIHSS≤4) | All | 64.9% | 71.0% | 70.7% | 69.8% | 0.06 |
| | Men | 66.8% | 72.4% | 72.1% | 70.1% | 0.44 |
| | Women | 59.4% | 67.5% | 67.2% | 69.1% | **0.02** |
| Moderate stroke (5-15) | All | 28.3% | 23.4% | 24.0% | 25.3% | 0.39 |
| | Men | 25.8% | 22.1% | 23.5% | 25.0% | 0.69 |
| | Women | 35.2% | 26.7% | 25.2% | 26.0% | **0.02** |
| Severe stroke (16-42) | All | 6.8% | 5.6% | 5.4% | 4.9% | **0.03** |
| | Men | 7.4% | 5.5% | 4.5% | 4.9% | **0.02** |
| | Women | 5.4% | 5.8% | 7.6% | 4.8% | 0.74 |
| **Reperfusion therapy** | | | | | | |
| IVT | All | 9.5% | 13.3% | 14.0% | 13.8% | .004 |
| | Men | 9.0% | 12.0% | 14.5% | 14.4% | <.001 |
| | Women | 10.7% | 16.8% | 12.8% | 12.4% | 0.57 |
| DTN≤ 45 min | All | 28.1% | 61.1% | 69.4% | 64.8% | < .001 |
| | Men | 27.0% | 63.6% | 73.9% | 65.8% | < .001 |
| | Women | 30.8% | 56.8% | 56.2% | 62.0% | 0.03 |
| DTN≤ 60 min | All | 56.2% | 84.2% | 86.3% | 81.5% | <.001 |
| | Men | 55.6% | 86.0% | 88.0% | 81.9% | .002 |
| | Women | 57.7% | 81.1% | 81.2% | 80.3% | 0.11 |
| Mechanical thrombectomy | All | 3.2% | 5.3% | 7.5% | 9.2% | <.001 |
| | Men | 3.6% | 4.6% | 6.6% | 9.0% | <.001 |
| | Women | 2.3% | 7.0% | 9.7% | 9.8% | <.001 |
| DTP≤ 90 min | All | 26.1% | 23.5% | 31.2% | 30.9% | 0.28 |
| | Men | 22.2% | 32.1% | 31.8% | 30.8% | 0.74 |
| | Women | 40.0% | 9.4% | 30.0% | 31.0% | 0.14 |
| **Discharge prescription** | | | | | | |
| DAPT on mild noncardiogenic stroke | All | 28.4% | 23.5% | 34.2% | 50.4% | <.001 |
| | Men | 29.3% | 23.7% | 36.1% | 53.0% | <.001 |
| | Women | 25.8% | 22.8% | 29.3% | 43.9% | <.001 |
| Warfarin on AF | All | 75.0% | 72.4% | 41.0% | 14.6% | <.001 |
| | Men | 68.4% | 72.2% | 36.8% | 8.7% | <.001 |
| | Women | 86.4% | 72.7% | 54.2% | 29.6% | <.001 |
| DOAC on AF | All | 0.0% | 4.6% | 30.0% | 56.2% | <.001 |
| | Men | 0.0% | 5.6% | 32.9% | 60.9% | <.001 |
| | Women | 0.0% | 3.0% | 20.8% | 44.4% | <.001 |
| Statin | All | 71.5% | 79.9% | 86.9% | 91.2% | <.001 |
| | Men | 73.1% | 81.9% | 88.4% | 92.5% | <.001 |
| | Women | 67.0% | 74.7% | 83.2% | 88.2% | <.001 |

* P-value by Cochran-Armitage trend test

AF, Atrial fibrillation; DTN, Door-to-needle time; DTP, Door-to-puncture time; IVT, Intravenous thrombolysis; MT, Mechanical thrombectomy; NIHSS, National Institution of Health Stroke Scale.

**Table S7**. Clinical Outcomes Stratified by Sex.

| Variables | | 2011-2013 (n=1753) | 2014-2016 (n=2055) | 2017-2019 (n=2249) | P for Trend* |
|---|---|---|---|---|---|
| **In-hospital Death** | All | 0.90% | 1.50% | 1.00% | 0.71 |
| | Men | 1.0% | 1.6% | 0.9% | 0.96 |
| | Women | 0.4% | 1.2% | 1.2% | 0.21 |
| **3-month mRS 0-1** | All | 67.2% | 67.8% | 68.6% | 0.37 |
| | Men | 68.5% | 68.7% | 68.3% | 0.90 |
| | Women | 63.7% | 65.7% | 69.2% | 0.06 |
| **3-month mRS 0-2** | All | 83.5% | 84.2% | 83.0% | 0.64 |
| | Men | 84.5% | 84.6% | 82.7% | 0.19 |
| | Women | 80.9% | 83.3% | 83.8% | 0.24 |
| **1-year Mortality** | All | 2.52 (1.77 - 3.28) | 2.93 (2.19 - 3.68) | 2.32 (1.69 - 2.95) | 0.50 |
| | Men | 2.57 (1.66 - 3.48) | 2.68 (1.82 - 3.53) | 2.00 (1.30 - 2.68) | 0.42 |
| | Women | 2.27 (0.87 - 3.66) | 3.22 (1.75 - 4.68) | 3.06 (1.69 - 4.40) | 0.78 |
| **1-year Cumulative Incidence of Recurrent Stroke** | All | 4.13 (3.17 - 5.09) | 3.70 (2.86 - 4.55) | 5.47 (4.51 - 6.44) | 0.04 |
| | Men | 4.06 (2.91 - 5.19) | 2.98 (2.06 - 3.88) | 5.71 (4.53 - 6.86 | 0.02 |
| | Women | 4.63 (2.62 - 6.60) | 5.32 (3.42 - 7.19) | 4.77 (3.06 - 6.45) | 0.95 |

* P-value by Mantel-Haenszel Chi-square test for ordinal variable, Cochran-Armitage trend test for categorical variables, linear contrast test in ANOVA for continuous variables, and linear contrast test in log-rank test for survival variables.

**Table S8**. Comparison between patients with and without 3 month mRS.

| Variables | With 3 month mRS (n=5809) | No 3 month mRS (n=248) | Total (n=6057) | *p-value** |
|---|---|---|---|---|
| Men | 71.6% | 70.6% | 71.6% | 0.72 |
| Mean age (SD), years | 43.1 (6.6) | 42.3 (7.3) | 43.0 (6.6) | 0.07 |
| **Risk factors** | | | | |
| Current smoking | 46.7% | 45.3% | 46.7% | 0.67 |
| Obesity (BMI ≥30 kg/m2) | 10.3% | 8.5% | 10.3% | 0.36 |
| Hypertension | 42.8% | 35.9% | 42.5% | 0.03 |
| Diabetes mellitus | 20.2% | 21.4% | 20.3% | 0.66 |
| Hyperlipidemia | 27.3% | 23.0% | 27.1% | 0.13 |
| Atrial fibrillation | 4.6% | 5.2% | 4.7% | 0.66 |
| **Ischemic stroke subtypes** | | | | |
| Large artery atherosclerosis | 30.5% | 31.5% | 30.5% | 0.75 |
| Small vessel occlusion | 22.2% | 21.8% | 22.2% | 0.88 |
| Cardioembolic stroke | 10.6% | 9.7% | 10.6% | 0.63 |
| Other determined etiology | 14.0% | 11.3% | 13.9% | 0.22 |
| Undetermined etiology | 22.8% | 25.8% | 22.9% | 0.24 |
| **Onset-to-door time** | | | | |
| Median FAT-to-door (IQR), h | 8.47 (2.12-32.78) | 7.27 (2.30-34.93) | 8.35 (2.12-32.85) | 0.52 |
| Median LKW-to-door (IQR), h | 11.68 (2.97-37.03) | 9.73 (3.04-36.70) | 11.63 (2.97-37.03) | 0.28 |
| **Initial Stroke severity** | | | | |
| Mean initial NIHSS score (SD) | 4.0 (5.0) | 5.3 (5.8) | 4.1 (5.1) | .002 |
| Median initial NIHSS score (IQR) | 2 (1-5) | 3 (1-7) | 2 (1-5) | |
| **Reperfusional therapy** | | | | |
| IVT rate | 13.8% | 12.5% | 13.8% | 0.56 |
| Median DNT (IQR), min | 38 (29-53) | 39 (29.5-49.25) | 38 (29-53) | 0.60 |
| DNT ≤60 min | 83.7% | 89.3% | 83.9% | 0.62 |
| MT rate | 7.4% | 9.3% | 7.5% | 0.28 |
| Median DPT (IQR), min | 110 (87-155) | 122 (92.75-145.75) | 110 (87-154) | 0.62 |
| DPT ≤90 min | 29.8% | 22.7% | 29.4% | 0.48 |
| **Discharge medications** | | | | |
| Dual antiplatelet therapy | 42.6% | 35.9% | 42.3% | 0.16 |
| Anticoagulation | 10.4% | 8.5% | 10.2% | 0.41 |
| Statin | 86.9% | 76.7% | 86.5% | <.001 |

* P-value by chi-square test for categorical variables and t-test for continuous variables.

FAT, first abnormal time; LKW, last known well; IVT, intravenous thrombolysis; DNT, door to needle time; MT, mechanical thrombectomy; DPT, door to puncture time.

**Table S9**. 3-months Functional Outcome and Mortality After Stroke According to Reperfusion Therapy.

| **Reperfusion Therapy** | **Outcomes** | **2011-2013 (n=1631)** | **2014-2016 (n=1980)** | **2017-2019 (n=2198)** | **P for trend*** |
|---|---|---|---|---|---|
| None (n= 4824) | Mean mRS (SD) | 2.14 (1.38) | 2.11 (1.27) | 2.15 (1.31) | 0.642 |
| | Median mRS (IQR) | 2 (1-3) | 2 (1-3) | 2 (1-3) | - |
| | mRS 0-1 | 70.4% | 71.3% | 71.7% | 0.420 |
| | mRS 0-2 | 85.9% | 87.3% | 86.0% | 0.948 |
| | Mortality (mRS = 6) | 1.9% | 1.2% | 1.6% | 0.508 |
| IVT (n=927) | Mean mRS (SD) | 2.65 (1.46) | 2.75 (1.65) | 2.67 (1.66) | 0.760 |
| | Median mRS (IQR) | 2 (1-4) | 2 (1-4) | 2 (1-4) | |
| | mRS 0-1 | 51.8% | 52.7% | 57.4% | 0.185 |
| | mRS 0-2 | 71.8% | 72.0% | 71.3% | 0.883 |
| | Mortality (mRS = 6) | 2.2% | 5.3% | 3.6% | 0.475 |
| MT (n=487) | Mean mRS (SD) | 3.29 (1.65) | 3.05 (1.93) | 3.12 (1.93) | 0.628 |
| | Median mRS (IQR) | 3 (2-4) | 3 (1-4) | 3 (1-4) | - |
| | mRS 0-1 | 33.7% | 47.7% | 48.7% | 0.036* |
| | mRS 0-2 | 57.0% | 67.1% | 62.9% | 0.528 |
| | Mortality (mRS = 6) | 5.8% | 10.7% | 8.6% | 0.621 |

* P-value by Cochran-Armitage trend test for categorical variables and ANOVA test for continuous variable.
IVT, Intravenous thrombolysis; MT, Mechanical thrombectomy.

**Table S10**. 3-months Functional Outcome and Mortality After Stroke According to Baseline Stroke Severity.

| **Initial Stroke Severity** | **Outcomes** | **2011-2013 (n=1631)** | **2014-2016 (n=1980)** | **2017-2019 (n=2198)** | **P for trend*** |
|---|---|---|---|---|---|
| Mild (NIHSS 0-4, n=4117) | Mean mRS (SD) | 1.82 (1.06) | 1.83 (1.01) | 1.91 (1.08) | 0.046 |
| | Median mRS (IQR) | 2 (1-2) | 2 (1-2) | 2 (1-2) | - |
| | mRS 0-1 | 79.7% | 80.9% | 79.2% | 0.672 |
| | mRS 0-2 | 93.8% | 94.2% | 91.5% | 0.015 |
| | Mortality (mRS = 6) | 0.80% | 0.69% | 0.64% | 0.605 |
| Moderate (NIHSS 5-15, n=1393) | Mean mRS (SD) | 2.96 (1.50) | 3.04 (1.55) | 2.91 (1.56) | 0.395 |
| | Median mRS (IQR) | 3 (2-4) | 3 (2-4) | 3 (2-4) | **-** |
| | mRS 0-1 | **41.1%** | **39.5%** | **47.1%** | **0.045*** |
| | mRS 0-2 | 66.2% | 64.5% | 68.6% | 0.387 |
| | Mortality (mRS = 6) | 3.17% | 3.25% | 2.81% | 0.728 |
| Severe (NIHSS 16-42, n=299) | Mean mRS (SD) | 4.64 (1.63) | 4.18 (1.92) | 4.26 (2.04) | 0.204 |
| | Median mRS (IQR) | 5 (4-6) | 4 (3-6) | 4 (2.75-6) | - |
| | mRS 0-1 | **13.2%** | **20.2%** | **25.0%** | **0.039*** |
| | mRS 0-2 | 22.0% | 38.5% | 33.7% | 0.094 |
| | Mortality (mRS = 6) | 10.2% | 15.5% | 14.6% | 0.374 |

* P-value by Cochran-Armitage trend test for categorical variables and ANOVA test for continuous variable.
NIHSS, National Institute of Health Stroke Scale, IVT, intravenous thrombolysis; MT, mechanical thrombectomy.

**Table S11**. Subgroup analysis of cumulative incidence of recurrent stroke stratified by ischemic stroke subtype.

| Ischemic Stroke Subtypes | 2011-2013 | 2014-2016 | 2017-2019 | p for trend |
|---|---|---|---|---|
| Large artery atherosclerosis | | | | 0.30 |
| recurrent stroke 30d | 2.62 (1.31-3.93) | 2.93 (1.60-4.27) | 1.60 (0.62-2.58) | |
| recurrent stroke 60d | 3.70 (2.15-5.25) | 3.27 (1.86-4.68) | 1.76 (0.73-2.79) | |
| recurrent stroke 365d | 5.01 (3.20-6.81) | 4.71 (3.00-6.41) | 3.30 (1.87-4.72) | |
| Small vessel occlusion | | | | 0.85 |
| recurrent stroke 30d | 1.02 (0.03-2.01) | 1.34 (0.27-2.40) | 1.46 (0.39-2.53) | |
| recurrent stroke 60d | 2.07 (0.65-3.48) | 1.57 (0.42-2.73) | 1.88 (0.66-3.09) | |
| recurrent stroke 365d | 3.45 (1.60-5.29) | 2.55 (1.06-4.03) | 3.41 (1.76-5.05) | |
| Cardioembolic stroke | | | | 0.02 |
| recurrent stroke 30d | 1.04 (0.00-2.48) | 0.44 (0.00-1.30) | 3.26 (0.88-5.64) | |
| recurrent stroke 60d | 1.61 (0.00-3.41) | 0.91 (0.00-2.17) | 4.72 (1.86-7.58) | |
| recurrent stroke 365d | 2.24 (0.06-4.41) | 2.50 (0.33-4.67) | 6.74 (3.33-10.16) | |
| Other determined etiology | | | | 0.20 |
| recurrent stroke 30d | 3.43 (0.73-6.13) | 2.29 (0.48-4.11) | 3.31 (1.54-5.09) | |
| recurrent stroke 60d | 5.85 (2.33-9.38) | 2.68 (0.72-4.65) | 5.40 (3.15-7.65) | |
| recurrent stroke 365d | 6.46 (2.76-10.16) | 6.82 (3.69-9.96) | 9.52 (6.56-12.49) | |
| Undetermined etiology | | | | 0.03 |
| recurrent stroke 30d | 1.88 (0.50-3.26) | 0.22 (0.00-0.64) | 3.82 (2.14-5.51) | |
| recurrent stroke 60d | 1.88 (0.50-3.26) | 0.88 (0.02-1.75) | 4.85 (2.95-6.74) | |
| recurrent stroke 365d | 3.35 (1.48-5.21) | 2.32 (0.90-3.74) | 6.52 (4.33-8.70) | |

**Figure S1**. Stroke Recurrence In Cardioembolic Stroke According to Discharge Medication (Warfarin vs. Others).

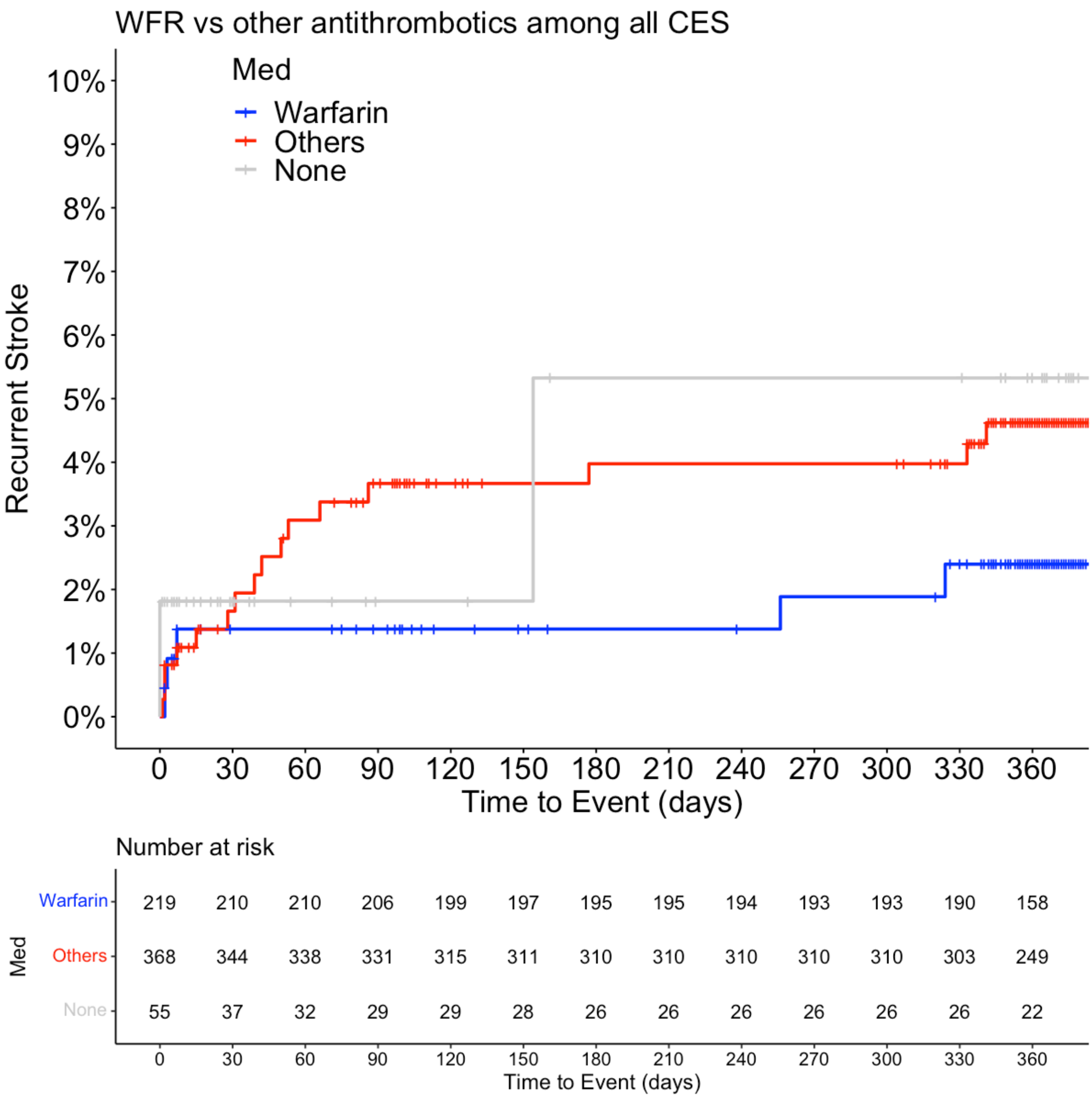

**Figure S2**. Stroke Recurrence In Other Determined Etiologies According to Discharge Medication (Warfarin vs. Others).

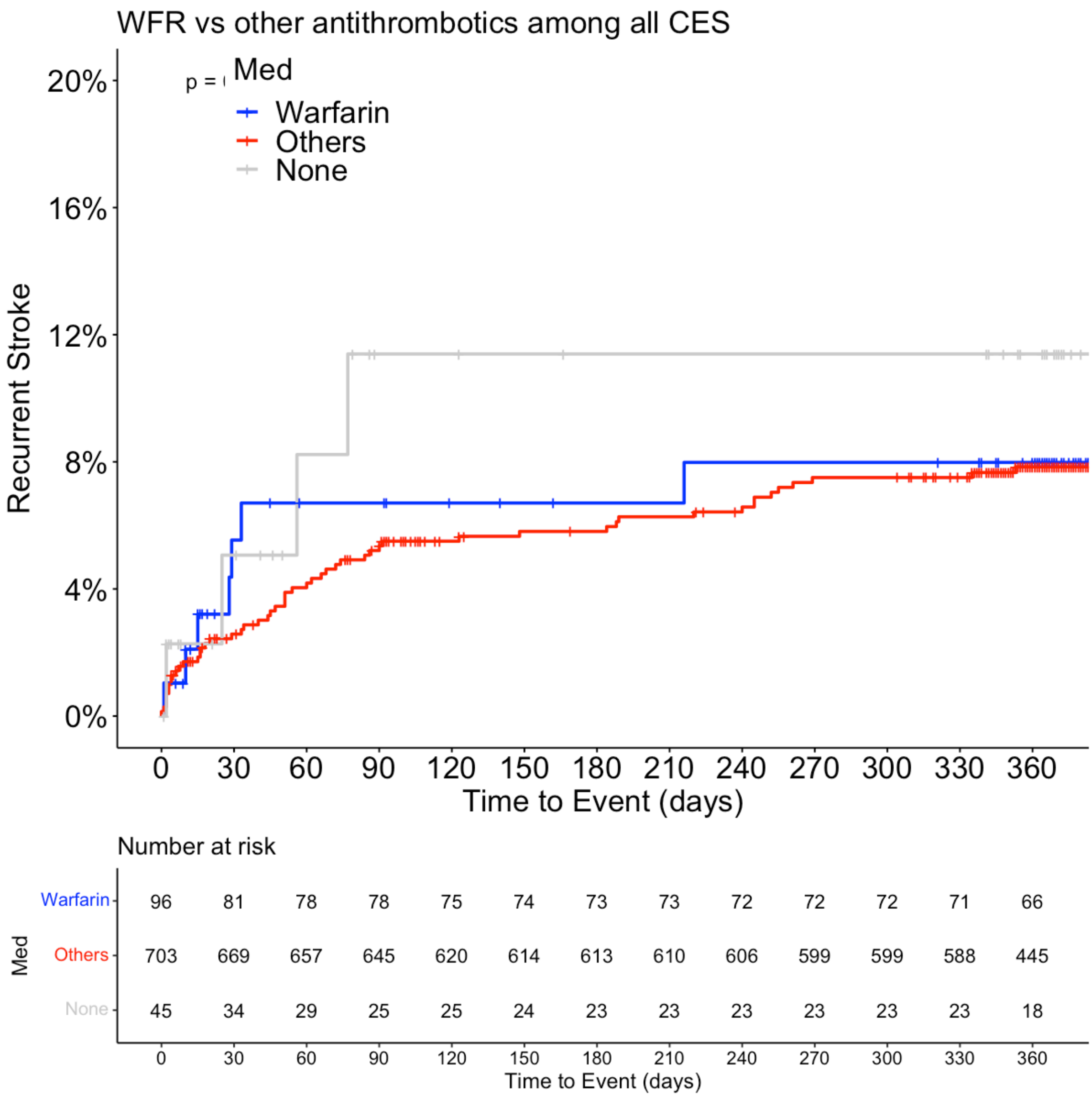
